# PCR assay to enhance global surveillance for SARS-CoV-2 variants of concern

**DOI:** 10.1101/2021.01.28.21250486

**Authors:** Chantal B.F. Vogels, Mallery I. Breban, Tara Alpert, Mary E. Petrone, Anne E. Watkins, Isabel M. Ott, Jaqueline Goes de Jesus, Ingra Morales Claro, Giulia Magalhães Ferreira, Myuki A.E. Crispim, Brazil-UK CADDE Genomic Network, Lavanya Singh, Houriiyah Tegally, Ugochukwu J. Anyaneji, NGS-SA, Emma B. Hodcroft, Christopher E. Mason, Gaurav Khullar, Jessica Metti, Joel T. Dudley, Matthew J. MacKay, Megan Nash, Jianhui Wang, Chen Liu, Pei Hui, Steven Murphy, Caleb Neal, Eva Laszlo, Marie L. Landry, Anthony Muyombwe, Randy Downing, Jafar Razeq, Tulio de Oliveira, Nuno R. Faria, Ester C. Sabino, Richard A. Neher, Joseph R. Fauver, Nathan D. Grubaugh

**Author notes:** Correspondence: Chantal B.F. Vogels, Joseph R. Fauver, Nathan D. Grubaugh. These authors contributed equally (Chantal B.F. Vogels, Mallery I. Breban). Senior authors (Joseph R. Fauver, Nathan D. Grubaugh).

## Abstract

With the emergence of SARS-CoV-2 variants that may increase transmissibility and/or cause escape from immune responses^1–3^, there is an urgent need for the targeted surveillance of circulating lineages. It was found that the B.1.1.7 (also 501Y.V1) variant first detected in the UK^4,5^ could be serendipitously detected by the ThermoFisher TaqPath COVID-19 PCR assay because a key deletion in these viruses, spike Δ69-70, would cause a “spike gene target failure” (SGTF) result. However, a SGTF result is not definitive for B.1.1.7, and this assay cannot detect other variants of concern that lack spike Δ69-70, such as B.1.351 (also 501Y.V2) detected in South Africa^6^ and P.1 (also 501Y.V3) recently detected in Brazil^7^. We identified a deletion in the ORF1a gene (ORF1a Δ3675-3677) in all three variants, which has not yet been widely detected in other SARS-CoV-2 lineages. Using ORF1a Δ3675-3677 as the primary target and spike Δ69-70 to differentiate, we designed and validated an open source PCR assay to detect SARS-CoV-2 variants of concern^8^. Our assay can be rapidly deployed in laboratories around the world to enhance surveillance for the local emergence spread of B.1.1.7, B.1.351, and P.1.

## Main

Broadly accessible and inexpensive surveillance methods are needed to track SARS-CoV-2 variants of concern around the world. While sequencing is the gold standard to identify circulating SARS-CoV-2 variants, routine genomic surveillance is not available in many locations primarily due to a lack of resources and expertise. In the current situation with the identification of the variants of concern B.1.1.7, B.1.351, and P.1, and with the likelihood that more will emerge, a lack of genomic surveillance leaves public health authorities with a patchy and skewed picture to inform decision making. The discovery of B.1.1.7 variants causing SGTF results when tested using the TaqPath PCR assay provided labs in the UK and throughout Europe with a ready-made, simple tool for tracking the frequencing of this variant^9,10^. As B.1.1.7 spread to other countries, TaqPath SGTF results were used as a front line screening tool for sequencing and an approximation for B.1.1.7 population frequency^11^. These findings highlight the usefulness of a PCR assay that produces distinctive results when targeting variants in virus genomes for both tracking and sequencing prioritization.

The TaqPath assay was not specifically designed for SARS-CoV-2 variant surveillance, and it has several limitations. The 6 nucleotide deletion in the spike gene at amino acid positions 69 and 70 (spike Δ69-70) that causes the TaqPath SGTF is also present in other SARS-CoV-2 lineages (**Fig. 1, Supplementary Table 1**), most notably Pango lineages B.1.258 detected throughout Europe and B.1.375 detected primarily in the US^12,13^, meaning that SGTF results are not definitive for B.1.1.7. Furthermore, too much focus on TaqPath SGTF results will leave blindspots for other emerging SARS-CoV-2 variants of concern that do not have spike Δ69-70. In particular, B.1.351 and P.1, which were recently discovered in South Africa and Brazil, respectively, may also be more transmissible and contain mutations that could help to evade immune responses^6,7,14^. For all of these reasons, a PCR assay specifically designed for variant surveillance would help to fill in many of the gaps about their distribution and frequency.

**Figure 1.**
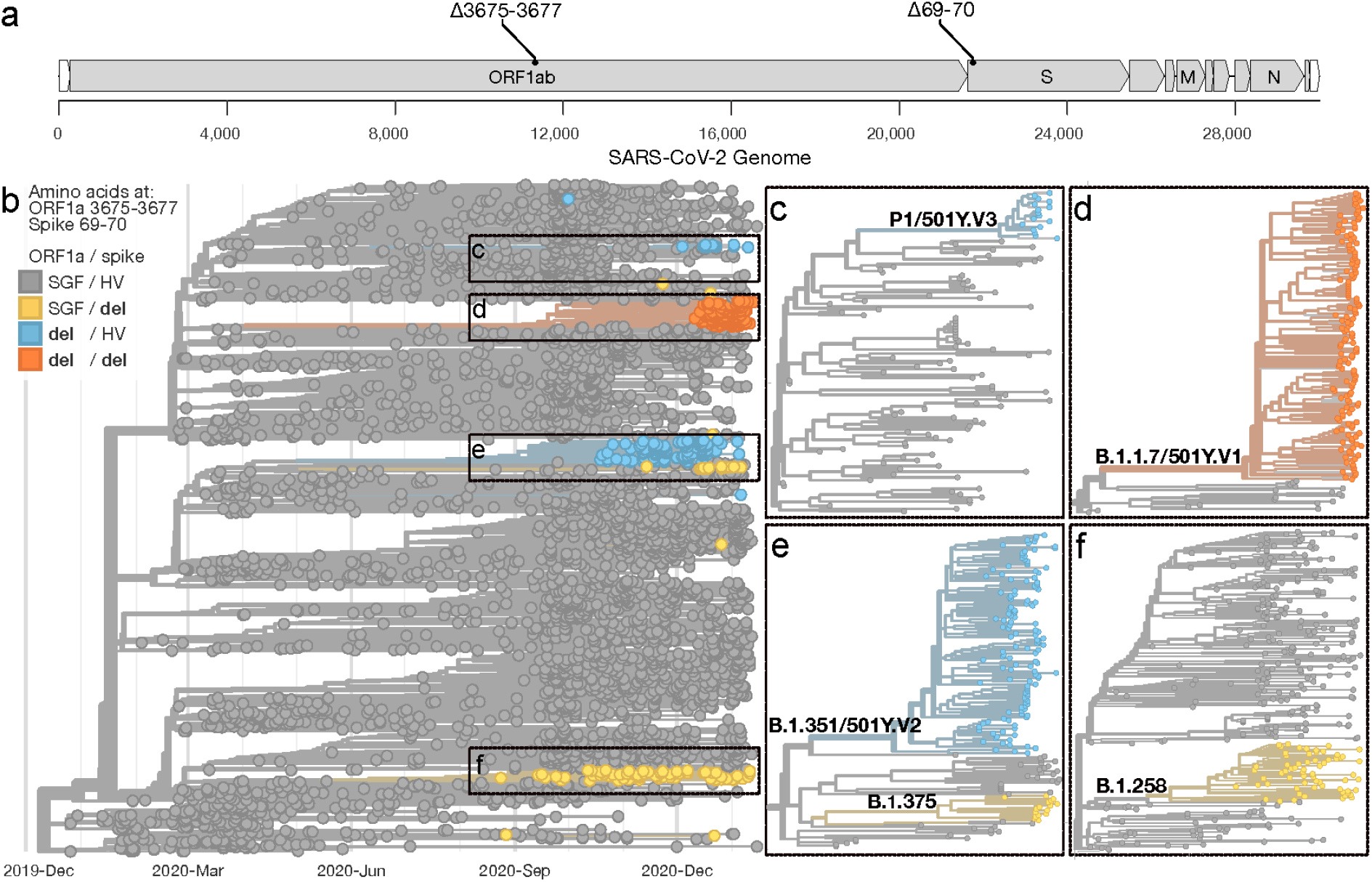
Identification of genome targets to differentiate between B.1.1.7, B.1.351, P.1, and other SARS-CoV-2 lineages. (**a**) Location on the SARS-CoV-2 genome where the targeted deletions in the ORF1a gene at amino acid positions 3675-3677 (Δ3675-3677) and the spike gene at amino acid positions 69-70 (Δ69-70) occur. (**b**) The Nextstrain ‘global build’ (nextstrain.org/ncov/global) accessed on 2021-01-22 showing the phylogenetic representation of 4,046 SARS-CoV-2 genomes colored by the presence of deletions at amino acid positions ORF1a 3575-3677 and spike 69-70. (**c-f**) Zooms of large SARS-CoV-2 clades, which include the variants of concern B.1.1.7, B.1.351, and P.1, containing one or both deletions. A list of SARS-CoV-2 genomes used in the analysis is available in **Source Data Fig. 1**.

We analyzed over 400,000 SARS-CoV-2 genomes on GISAID and used custom Nextstrain builds^15^ to identify that a 9 nucleotide deletion in the ORF1a gene at amino acid positions 3675-3677 (ORF1a Δ3675-3677) occurs in the B.1.1.7, B.1.351, and P.1 variants, but is only found in 0.03% (103/377,011) of all other genomes (**Fig. 1, Supplementary Table 1**). Within the B.1.351 lineage, however, 18.4% of the sequences do not have ORF1a Δ3675-3677 (**Supplementary Table 1**, not shown in **Fig. 1e**). By designing a PCR assay that targets both ORF1a Δ3675-3677 and spike Δ69-70 (**Fig. 1a**), we can detect most viruses from all three current variants of concern (ORF1a results, **Fig. 1b-e**), differentiate B.1.1.7 (ORF1a and spike results, **Fig. 1d-f**), and provide results similar to TaqPath SGTF to compare datasets (spike results).

To create a multiplexed RT-qPCR screening assay for the B.1.1.7, B.1.351, and P.1 variants, we designed two sets of primers that flank each of ORF1a Δ3675-3677 and spike Δ69-70 and probes specific to the undeleted “wildtype” sequences. As a control, we included the CDC N1 (nucleocapsid) primer and probe set that will detect both the wildtype and variant viruses^16^. As designed, testing SARS-CoV-2 RNA that contains ORF1a Δ3675-3677 and/or spike Δ69-70 will generate undetected cycle threshold (Ct) values with the specific PCR target sets as the probes cannot anneal to the deleted sequences, but will have “positive” N1 Ct values. This configuration ensures that target failures are likely due to the presence of deletions and that there is sufficient virus RNA for sequencing confirmation. Our RT-qPCR conditions are highly similar to our previously published SARS-CoV-2 multiplex assay^17^, and a detailed protocol is openly available^8^.

We evaluated the analytical sensitivity of our multiplexed RT-qPCR assay using synthetic RNA designed based on the original Wuhan-Hu-1 sequence and a B.1.1.7 sequence (England/205041766/2020). As the B.1.1.7 sequence contains both ORF1a Δ3675-3677 and spike Δ69-70 and the Wuhan-Hu-1 sequence contains neither deletion, using these RNAs allows us to fully evaluate the designed primer and probe sets. We tested a two-fold dilution series from 100 copies/µL to 1 copy/µL for both RNA controls in triplicate (**Table 1**). Using the Wuhan-Hu-1 RNA, we found similar detection (within 1 Ct) across all three N1, ORF1a, and spike targets, and all three could detect virus RNA at our lowest concentration of 1 copy/µL, indicating that our primer and probes sets were efficiently designed. Using the B.1.1.7 RNA, we again could detect the RNA down to 1 copy/µL with the N1 set, but did not detect any concentration of the virus RNA with the ORF1a and spike sets, confirming the expected “target failure” signature when testing viruses containing both ORF1a Δ3675-3677 and spike Δ69-70. Overall, our PCR screening assay could easily differentiate between SARS-CoV-2 RNA with and without the ORF1a and spike deletions by comparing the Ct values to the N1 control.

**Table 1:** Analytical sensitivity of the multiplexed RT-qPCR assay to screen for variants of concern using primer/probe sets targeting key deletions. Listed are cycle threshold (Ct) values for the three primer-probe sets targeting the SARS-CoV-2 nucleocapsid (N1-FAM), ORF1a 3675-3766 deletion (ORF1a-Cy5), and spike 69-70 deletion (Spike-HEX). Two-fold dilutions of synthetic control RNA (Wuhan-Hu-1 and B.1.1.7) were tested in triplicate.

| RNA | Concentration | N1-FAM |  |  | ORF1a-Cy5 |  |  | Spike-HEX |  |  |
| --- | --- | --- | --- | --- | --- | --- | --- | --- | --- | --- |
| Wuhan-Hu-1 | 100 copies/μL | 30.4 | 30.5 | 30.4 | 31.1 | 31.2 | 31.1 | 30.8 | 30.8 | 30.9 |
| | 50 copies/μL | 31.5 | 31.5 | 31.4 | 32.1 | 32.1 | 32.0 | 31.9 | 31.6 | 31.7 |
| | 25 copies/μL | 32.3 | 32.7 | 32.7 | 33.0 | 33.1 | 33.2 | 32.8 | 32.8 | 33.0 |
| | 12 copies/μL | 33.3 | 33.7 | 33.3 | 34.1 | 34.2 | 34.2 | 34.0 | 33.6 | 34.1 |
| | 6 copies/μL | 34.5 | 34.3 | 34.9 | 35.1 | 35.2 | 35.4 | 35.1 | 34.3 | 35.6 |
| | 3 copies/μL | 35.0 | 37.0 | 36.1 | 35.6 | 36.6 | 36.8 | 35.4 | 36.1 | 35.8 |
| | 1 copy/μL | 37.0 | 37.0 | 35.7 | 37.2 | 36.7 | 37.0 | 36.4 | 36.4 | 36.3 |
| B.1.1.7 | 100 copies/μL | 29.1 | 29.2 | 29.2 | ND | ND | ND | ND | ND | ND |
| | 50 copies/μL | 30.1 | 30.2 | 30.2 | ND | ND | ND | ND | ND | ND |
| | 25 copies/μL | 31.1 | 31.2 | 31.2 | ND | ND | ND | ND | ND | ND |
| | 12 copies/μL | 32.5 | 32.2 | 32.1 | ND | ND | ND | ND | ND | ND |
| | 6 copies/μL | 33.0 | 33.2 | 33.1 | ND | ND | ND | ND | ND | ND |
| | 3 copies/μL | 33.7 | 34.6 | 34.0 | ND | ND | ND | ND | ND | ND |
| | 1 copy/μL | 35.0 | 35.2 | 35.3 | ND | ND | ND | ND | ND | ND |

Wuhan-Hu-1 = Twist synthetic RNA control 2; B.1.1.7 = Twist synthetic RNA control 14; N1-FAM = CDC N1 primer-probe set targeting the nucleocapsid with FAM fluorophore<sup>16</sup>; ORF1a-Cy5 = Yale primer-probe set targeting the ORF1a gene 3675-3677 deletion with Cy5 fluorophore<sup>8</sup>; Spike-HEX = Yale primer-probe set targeting the spike gene 69-70 deletion with HEX fluorophore<sup>8</sup>.

Next, we validated our multiplex RT-qPCR variant screening assay using 376 known COVID-19 clinical samples that we have previously sequenced in our laboratories in the US, Brazil, and South Africa (**Fig. 2, Source Data Fig 2**). We tested 210 samples from SARS-CoV-2 lineages without either ORF1a Δ3675-3677 and spike Δ69-70 (classified as “other” lineage), 68 samples with both ORF1a Δ3675-3677 and spike Δ69-70 deletions, 51 samples with only the ORF1a Δ3675-3677 deletion, and 47 samples with only the spike Δ69-70 deletion. Of the samples without both deletions (expected outcome = detection with all three primer/probe sets), 2.9% (6/210) were P.2 (variant of interest [VOI]) and 97.1% (204/210) were other lineages not of current interest (**Fig. 2a**). Of the samples with both ORF1a Δ3675-3677 and spike Δ69-70 deletions (expected outcome = target failure with both the ORF1a and spike sets), 97.1% (67/69) were B.1.1.7 (VOC) and 2.9% (2/69) were B.1.525 (VOI; **Fig. 2b**). Of the samples with ORF1a Δ3675-367, 47.1% (24/51) were B.1.351 (VOC), 31.4% (16/51) were P.1 (VOC), 7.8% (4/51) were B.1.526 (VOI), and 13.7% (7/51) were other lineages not of current interest (**Fig. 2c**). Of the samples with only the spike Δ69-70 deletion, 87.2% (41/47) were B.1.375 and 12.8% (6/47) were other lineages not of current interest (**Fig. 2d**). Importantly, unlike the TaqPath assay SGTF results, we could differentiate between B.1.1.7 and other variants that only have the spike deletion, such as B.1.375 that is not currently a variant of concern/interest.

**Figure 2.**
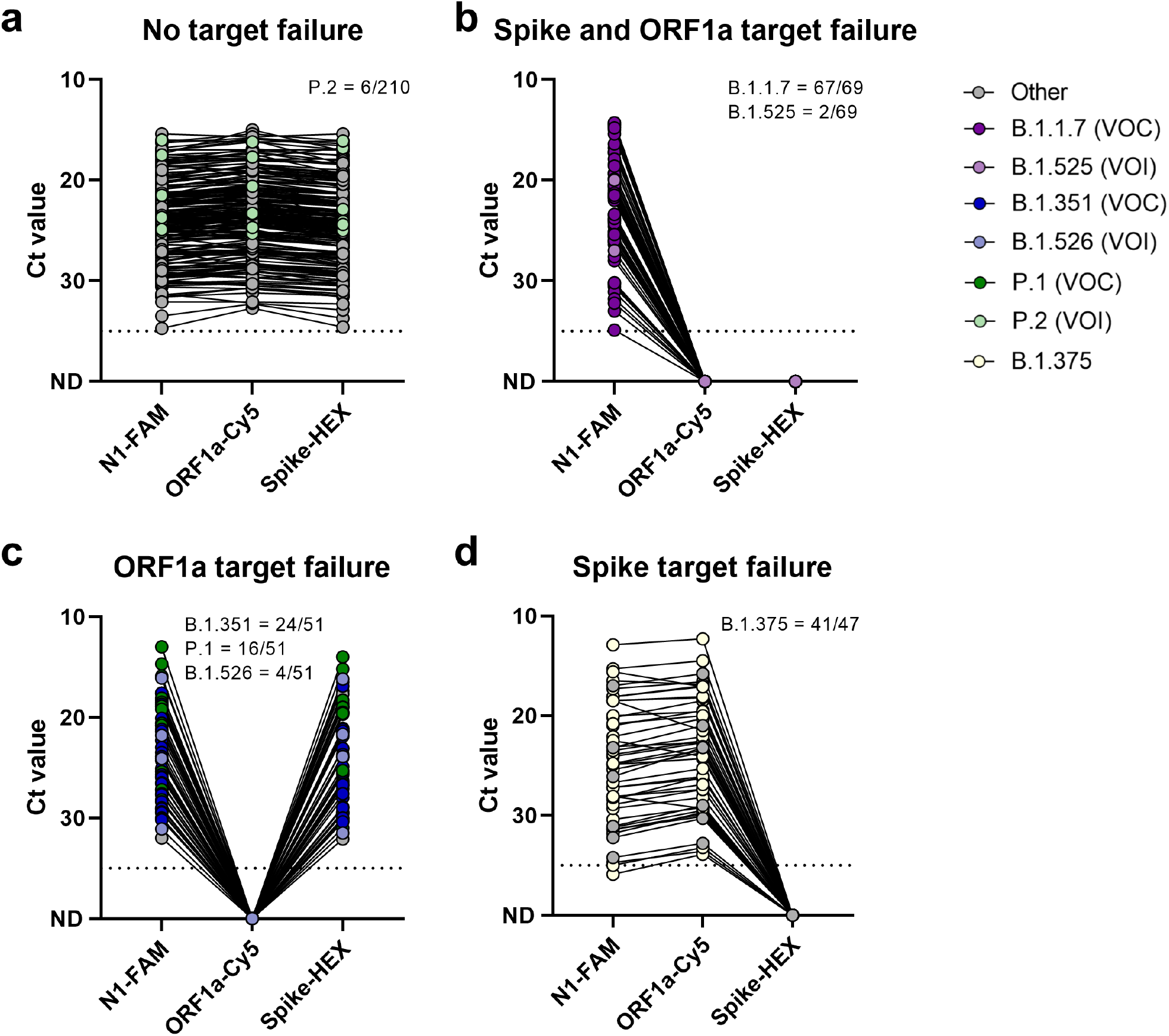
ORF1a and spike target failure used to differentiate between SARS-CoV-2 variants of concern B.1.1.7, B.1.351, P.1, and other lineages currently not of concern. (**a**) Other lineages without target failure are detected by all three targets of the multiplex RT-qPCR assay. (**b**) Double target failure indicates the presence of the ORF1a Δ3675-3677 and spike Δ69-70 deletions and can be used to identify potential B.1.1.7 variants. (**c**) ORF1a target failure indicates the presence of the ORF1a Δ3675-3677 deletion, present in B.1.351 and P.1 variants of concern. (**d**) Spike target failure indicates presence of the spike Δ69-70 deletion, which is present in various lineages, including B.1.375. Shown are the Ct values for the N1, ORF1a, and spike primer-probe sets, with lines connecting Ct values obtained with the three sets for the same specimen. The dotted line indicates the limit of detection. VOC = variant of concern, VOI = variant of interest. Data used to make this figure can be found in **Source Data Fig 2**.

When splitting up our findings by country, we tested a total of 305 samples from the US, 23 from Brazil, and 48 from South Africa. Within the US, the majority of samples were from Connecticut and fewer samples from the states of Arizona, Arkansas, Illinois, Massachusetts, North Carolina, Tennessee, Texas, Utah, and Wisconsin. Of the samples without target failure, 100% (180/180) samples belonged to other lineages not of current interest. Of the samples with double target failure, 97.1% (67/69) were B.1.1.7 (VOC) and 2.9% (2/69) were B.1.525 (VOI). Of the samples with ORF1a target failure, 40% (4/10) were B.1.526 (VOI) and 60% (6/10) were other lineages. Of the samples with spike target failure, 87.2% (41/47) were B.1.375 and 12.8% (6/47) were other lineages. Thus, if we used ORF1a target failure (with or without spike target failure) to identify high priority samples for sequencing, we would have confirmed 100% of the B.1.1.7 samples and detected two new variants of interest (B.1.525 and B.1.526, both containing the E484K mutation)^18^ while triaging 74% (207/305) of the samples.

In Brazil, we tested samples from the cities of São Paulo and Manaus. Of the samples without target failure, 85.7% (6/7) were P.2 (VOI) and 14.3% (1/7) were other lineages. Of the samples with ORF1a target failure, 100% (16/16) were P.1 (VOC). Within South Africa, samples were tested from the KwaZulu-Natal province. Of the samples without target failure, 100% (23/23) samples belonged to other lineages. Of the samples with ORF1a target failure, 96% (24/25) were B.1.351 (VOC) and 4% (1/25) belonged to other lineages. Thus, our clinical results demonstrate how our multiplex RT-qPCR assay can detect potential SARS-CoV-2 variants of concern and/or interest from a variety of settings to prioritize samples for sequencing.

There are some limitations to our study as presented here. First, we initially observed autofluorescence of the N1 primer-probe set when testing negative template controls. By lowering the N1 primers and probe concentrations to 200 nM and 100 nM per reaction, respectively, we have reduced autofluorescence to levels above our threshold of Ct 35 (autofluorescence detected in 3/48 reactions with average Ct 39.6). Importantly, this PCR assay should only be used to screen known SARS-CoV-2 positive clinical samples for the presence of key deletions found in variants of concern (where autofluorescence will not be a factor), and it should not be used as a primary clinical diagnostic. We also suggest using a N1 threshold Ct of 30-35 for calling target failures in the ORF1a and spike sets and performing whole genome sequencing to confirm the identity of variants. For instance, the threshold for the system used in Brazil was lowered to Ct 30 as there did not seem to be consistent detection of all three sets above that threshold. Thus, the threshold may differ between used RT-qPCR kits and instruments and needs to be determined by individual laboratories.

Second, our assay will not be 100% sensitive and/or specific to all variants of concern. For instance, there is a monophyletic clade within the B.1.351 lineage that has ORF1a Δ3675-3677 filled back in, perhaps due to recombination with viruses that did not have the deletion. Moreover, we detected other virus lineages with double target failure (notably B.1.525) and ORF1a target failure (notably B.1.526), which are not currently variants of concern. B.1.525 and B.1.526, however, are of interest as they do contain the E484K and other key spike mutations that require further evaluation of their implications for transmissibility and immune escape^18^. These examples demonstrate that continuous monitoring of the lineages that contain the ORF1a Δ3675-3677 and spike Δ69-70 deletions will be necessary to ensure the local effectiveness of our assay.

The rapid emergence of the SARS-CoV-2 variants of concern necessitates an immediate roll out of surveillance tools. Although whole genome sequencing is required to definitively identify specific variants, resource and capacity constraints can limit the number of samples that can be sequenced. The ThermoFisher TaqPath assay has demonstrated the value of PCR for variant surveillance, but it is limited to B.1.1.7 and cannot differentiate between other viruses containing spike Δ69-70. By targeting two different large nucleotide deletions, ORF1a Δ3675-3677 and spike Δ69-70, we demonstrate that our multiplex PCR can rapidly screen for B.1.1.7, B.1.351, and P.1 variants, detect other variants of interest, and differentiate between most non-variants of concern. Thus, our multiplex RT-qPCR variant screening assay can be used to prioritize samples for sequencing and as a surveillance tool to help monitor the distribution and population frequency of suspected variants.

## Methods

### Ethics

#### United States

The Institutional Review Board from the Yale University Human Research Protection Program determined that the RT-qPCR testing and sequencing of de-identified remnant COVID-19 clinical samples conducted in this study is not research involving human subjects (IRB Protocol ID: 2000028599).

#### Brazil

Sample collection and genetic characterization was approved under the Brazilian National IRB (CONEP) CAAE 30101720.1.0000.0068.

#### South Africa

We used de-identified remnant nasopharyngeal and oropharyngeal swab samples from patients testing positive for SARS-CoV-2 by RT–qPCR from public health and private medical diagnostics laboratories in South Africa. The project was approved by University of KwaZulu-Natal Biomedical Research Ethics Committee (protocol reference no. BREC/00001195/2020; project title: COVID-19 transmission and natural history in KwaZulu-Natal, South Africa: epidemiological investigation to guide prevention and clinical care). Individual participant consent was not required for the genomic surveillance. This requirement was waived by the Research Ethics Committees.

The sample IDs displayed in **Source Data Fig 2** are not known outside the research groups and cannot be used to re-identify any subject.

### Analysis of public SARS-CoV-2 genomes

All available SARS-CoV-2 data (402,899 genomes) were downloaded on 2021-01-22 from GISAID and evaluated for the presence of ORF1a Δ3675-3677 and spike Δ69-70. Phylogenetic analysis of a subset of 4,046 SARS-CoV-2 genomes was performed using Nextstrain^15^, downsampled as shown using the “global build” on 2021-01-22 (https://nextstrain.org/ncov/global). A list of SARS-CoV-2 genomes used in the analysis is available in **Source Data Fig. 1**.

### Multiplex RT-qPCR with probes

A detailed protocol of our multiplexed RT-qPCR to screen for SARS-COV-2 B.1.1.7, B.1.351, and P.1 variants of concern can be found on protocols.io^8^. In brief, our multiplex RT-qPCR assay consists of the CDC N1^16^, and the newly designed ORF1a Δ3675-3677 and spike Δ69-70 primer-probe sets (**Supplementary Table 2**). We used the NEB Luna universal probe one step RT-qPCR kit with 200 nM of N1 primers, 100 nM of N1 probe, 400 nM of the ORF1a and spike primers, 200 nM of ORF1a and spike probes, and 5 µL of nucleic acid in a total reaction volume of 20 µL. Thermocycler conditions were reverse transcription for 10 minutes at 55°C, initial denaturation for 1 minute at 95°C, followed by 40 cycles of 10 seconds at 95°C and 30 seconds at 55°C. During initial validation we ran the PCR for 45 cycles. Differentiation between variants of concern is based on target failure of the ORF1a and/or spike primer-probe sets (**Supplementary Table 3**).

### Limit of detection

We used Twist synthetic SARS-CoV-2 RNA controls 2 (Genbank ID: MN908947.3; GISAID ID: Wuhan-Hu-1) and control 14 (Genbank ID: EPI_ISL_710528; GISAID ID: England/205041766/2020) to determine the limit of detection of the screening RT-qPCR assay. We tested a two-fold dilution series from 100 copies/µL to 1 copy/µL for both RNA controls in triplicate, and confirmed the lowest concentration that was detected in all three replicates by 20 additional replicates.

### Validation and sequence confirmation

#### United States

We validated our approach using known SARS-CoV-2 positive clinical samples. Briefly, we extracted nucleic acid from 300 µL viral transport medium from nasopharyngeal swabs and eluted in 75 µL using the MagMAX viral/pathogen nucleic acid isolation kit (ThermoFisher Scientific). Extracted nucleic acid was tested by our multiplexed RT-qPCR assay and then sequenced using a slightly modified ARTIC Network nCoV-2019 sequencing protocol for the Oxford Nanopore MinION^19,20^. These modifications include extending incubation periods of ligation reactions and including a bead-based clean-up step following dA-tailing. MinION sequencing runs were monitored using RAMPART^21^. Consensus sequences were generated using the ARTIC Network bioinformatics pipeline and lineages were assigned using Pangolin v.2.0^22,23^. GISAID accession numbers for all SARS-CoV-2 genomes used to validate our approach are listed in **Source Data Fig 2**.

#### Brazil

To validate the detection of the P.1 lineage, we selected 23 samples (16 P.1 and 7 others) that had been sequenced using the ARTIC protocol on the MinION sequencing platform, as previously described^7,24^. In brief, viral RNA was isolated from RT-PCR positive samples using QIAamp Viral RNA Mini kit (Qiagen), following the manufacturer’s instructions. cDNA was synthesized with random hexamers and the Protoscript II First Strand cDNA synthesis Kit (New England Biolabs). Whole-genome multiplex-PCR amplification was then conducted using the ARTIC network SARS-CoV-2 V3 primer scheme with the Q5 High-Fidelity DNA polymerase (New England Biolabs). Multiplex-PCR products were purified by using AmpureXP beads (Beckman Coulter) and quantification was carried out using the Qubit dsDNA High Sensitivity assay on the Qubit 3.0 (Life Technologies). Samples were then normalised in an equimolar proportion of 10 ng per sample. After end repair and dA tailing, DNA fragments were barcoded using the EXP-NBD104 (1–12) and EXP-NBD114 (13–24) Native Barcoding Kits (Oxford Nanopore Technologies). Barcoded samples were pooled together and sequencing adapter ligation was performed using the SQK-LSK 109 Kit (Oxford Nanopore Technologies). Sequencing libraries were loaded onto an R9.4.1 flow-cell (Oxford NanoporeTechnologies) and sequenced using MinKNOW version 20.10.3 (Oxford Nanopore Technologies). We tested RNA from sequenced samples with the multiplex RT-qPCR as described above, using the Applied Biosystems 7500 real-time PCR machine and a lowered threshold of Ct 30.

#### South Africa

We extracted nucleic acid using the Chemagic 360 (PerkinElmer). Briefly, 200 μl of viral transport medium from each swab sample was extracted and eluted in 100 μl using the Viral NA / gDNA kit. Complementary DNA (cDNA) synthesis, PCR, whole genome sequencing and genome assembly was done as previously described in detail using the ARCTIC protocol^6,19^. Out of the sequenced samples, we selected 24 B.1.351 samples and 24 samples belonging to other lineages to validate the RT-qPCR assay. We adapted the protocol by using the TaqPath 1-Step multiplex master mix (ThermoFisher Scientific) with 5 μl of extracted nucleic acid in a total reaction volume of 20 μl. Samples were amplified using the QuantStudio 7 Flex Real-Time PCR System using the following PCR conditions: UNG incubation for 2 minutes at 25°C, reverse transcription for 15 minutes at 50°C, polymerase activation for 2 minutes at 95°C, followed by 40 cycles of amplification at 95°C for 3 seconds and 55°C for 30 seconds.

## Data Availability

Genomic data are available on GISAID (see Source Data Fig 2 for accession numbers). All RT-qPCR data are included in this article, supplementary files, and source data.

https://www.gisaid.org/

## Data availability

Genomic data are available on GISAID (see **Source Data Fig 2** for accession numbers). All RT-qPCR data are included in this article, supplementary files, and source data.

## Acknowledgements

We thank A. Brito, C. Chiu, A. Lauring, A. Altajar, D. Comstock, P. Jack, S. Taylor, and V. Parsons for diagnostic samples, clinical support, and/or discussion. A list of acknowledgements for the SARS-CoV-2 data used in Fig. 1 can be found in the **Source Data Fig. 1**. This work was funded by CTSA Grant Number TL1 TR001864 (TA and MEP), Wellcome Trust and Royal Society Sir Henry Dale Fellowship (204311/Z/16/Z), a Medical Research Council-São Paulo Research Foundation CADDE partnership award (MR/S0195/1 and FAPESP 18/14389-0), Fast Grant from Emergent Ventures at the Mercatus Center at George Mason University (NDG), and CDC Contract # 75D30120C09570 (NDG).

## Author information

### Affiliations

**Department of Epidemiology of Microbial Diseases, Yale School of Public Health, New Haven, CT 06510, USA**

Chantal B.F. Vogels, Mallery Breban, Tara Alpert, Mary E. Petrone, Anne E. Watkins, Isabel M. Ott, Joseph R. Fauver, Nathan D. Grubaugh

**Department of Ecology and Evolutionary Biology, Yale University, New Haven, CT 06510, USA**

Nathan D. Grubaugh

**Departamento de Molestias Infecciosas e Parasitarias and Instituto de Medicina Tropical da Faculdade de Medicina da Universidade de São Paulo, São Paulo, Brazil**

Jaqueline Goes de Jesus, Ingra Morales Claro, Giulia Magalhães Ferreira, Nuno R. Faria, Ester C. Sabino

**Laboratório de Virologia, Instituto de Ciências Biomédicas, Universidade Federal de Uberlândia, Uberlândia, MG, Brazil**

Giulia Magalhães Ferreira

**Fundação Hospitalar de Hematologia e Hemoterapia do Amazonas, Manaus, Brazil**

Myuki A.E. Crispim

**KwaZulu-Natal Research Innovation and Sequencing Platform (KRISP), School of Laboratory**

**Medicine & Medical Sciences, University of KwaZulu-Natal, Durban, South Africa**

Lavanya Singh, Houriiyah Tegally, Ugochukwu J. Anyaneji, Tulio de Oliveira

**Institute of Social and Preventive Medicine, University of Bern, Bern, Switzerland**

Emma B. Hodcroft

**Tempus Labs, Chicago, IL 60654, USA**

Christopher E. Mason, Gaurav Khullar, Jessica Metti, Joel T. Dudley, Matthew J. MacKay, Megan Nash

**Department of Pathology, Yale University School of Medicine, New Haven, CT 06510, USA**

Jianhui Wang, Chen Liu, Pei Hui

**Murphy Medical Associates, Greenwich, CT 06614, USA**

Steven Murphy, Caleb Neal, Eva Laszlo

**Departments of Laboratory Medicine and Medicine, Yale School of Medicine, New Haven, CT 06510, USA**

Marie L. Landry

**Connecticut State Department of Public Health, Rocky Hill, CT 06067, USA**

Anthony Muyombwe, Randy Downing, Jafar Razeq

**MRC Centre for Global Infectious Disease Analysis, J-IDEA, Imperial College London, London, UK**

Nuno R. Faria

**Department of Zoology, University of Oxford, Oxford, UK**

Nuno R. Faria

**Biozentrum, University of Basel, 4056 Basel, Switzerland**

Richard A. Neher

**Swiss Institute of Bioinformatics, 1015 Lausanne, Switzerland**

Richard A. Neher

### Brazil-UK CADDE Genomic Network

Flavia Cristina da Silva Sales, Mariana Severo Ramundo, Darlan S. Candido, Camila Alves Maia Silva, Mariana Cardoso de Pinho, Thais de Moura Coletti, Pâmela dos Santos Andrade, Leandro Menezes de Souza, Esmênia Coelho Rocha, Ana Carolina Gomes Jardim, Erika Manuli, Nelson Gaburo Jr, Celso Granato, José Eduardo Levi, Silvia Costa, William Marciel de Souza, Maria Anice Salum, Rafael Pereira, Andreza de Souza, Lucy E. Matkin, Mauricio L. Nogueria, Anna Sara Levin, Philippe Mayaud, Neal Alexander, Renato Souza, Andre Luis Acosta, Carlos Prete, Joshua Quick, Oliver Brady, Janey Messina, Moritz Kraemer, Nelson da Cruz Gouveia, Izabel Oliva, Marcilio de Souza, Carolina Lazari, Cecila Salete Alencar, Julien Thézé, Lewis Buss, Leonardo Araujo, Mariana S. Cunha, Nicholas J Loman, Oliver G. Pybus, Renato S. Aguiar.

### Network for Genomic Surveillance in South Africa (NGS-SA)

Eduan Wilkinson, Nokukhanya Msomi, Arash Iranzadeh, Vagner Fonseca, Deelan Doolabh, Emmanuel James San, Koleka Mlisana, Anne von Gottberg, Sibongile Walaza, Mushal Allam, Arshad Ismail, Thabo Mohale, Allison J. Glass, Susan Engelbrecht, Gert Van Zyl, Wolfgang Preiser, Francesco Petruccione, Alex Sigal, Diana Hardie, Gert Marais, Marvin Hsiao, Stephen Korsman, Mary-Ann Davies, Lynn Tyers, Innocent Mudau, Denis York, Caroline Maslo, Dominique Goedhals, Shareef Abrahams, Oluwakemi Laguda-Akingba, Arghavan Alisoltani-Dehkordi, Adam Godzik, Constantinos Kurt Wibmer, Bryan Trevor Sewell, José Lourenço, Sergei L. Kosakovsky Pond, Steven Weaver, Marta Giovanetti, Luiz Carlos Junior Alcantara, Darren Martin, Jinal N. Bhiman, Carolyn Williamson

### Contributions

CBFV, RAN, JRF, and NDG designed the study; CEM, GK, JM, JTD, MJM, MN, JW, CL, PH, SM, CN, EL, MLL, AM, RD, and JR collected and provided clinical samples; CBFV, MIB, TA, MEP, AEW, IMO, EBH, JGJ, IMC, GMF, MAEC, B-UCGN, NRF, ECS, LS, HT, UJA, NGS-SA, TO, RAN, JRF, and NDG collected and analyzed data; JRF and NDG supervised the project; CBFV and NDG wrote and edited the manuscript; all authors read and approved the final manuscript.

## Source Data

**Source Data Fig. 1:** Supporting data for Fig. 1.

**Source Data Fig. 2:** Supporting data for Fig. 2.

## Supplementary information

**Supplementary Table 1:** Summary of SARS-CoV-2 genomes with the ORF1a 3675-3677 and/or Spike 69-70 deletions (GISAID on 2020-01-21).

| Clade | Total sequences | ORF1a: Δ3675-3677 | Spike: Δ69-70 | % ORF1a: Δ3675-3677 | % Spike: Δ69-70 |
| --- | --- | --- | --- | --- | --- |
| 19A | 16,034 | 5 | 9 | 0.0% | 0.1% |
| 19B | 8,869 | 1 | 4 | 0.0% | 0.0% |
| 20A | 81,139 | 22 | 6,494 | 0.0% | 8.0% |
| 20A.EU2 | 10,380 | 5 | 13 | 0.0% | 0.1% |
| 20B | 95,315 | 21 | 1,366 | 0.0% | 1.4% |
| 20C | 45,041 | 43 | 274 | 0.1% | 0.6% |
| 20D | 4,804 | 0 | 4 | 0.0% | 0.1% |
| 20E.EU1 | 91,540 | 2 | 7 | 0.0% | 0.0% |
| 20F | 12,613 | 4 | 0 | 0.0% | 0.0% |
| 20G | 11,276 | 0 | 22 | 0.0% | 0.2% |
| B.1.351 | 582 | 474 | 0 | 81.4% | 0.0% |
| B.1.1.7 | 25,262 | 25,178 | 25,142 | 99.7% | 99.5% |
| P.1 | 34 | 34 | 0 | 100.0% | 0.0% |

**Supplementary Table 2:**
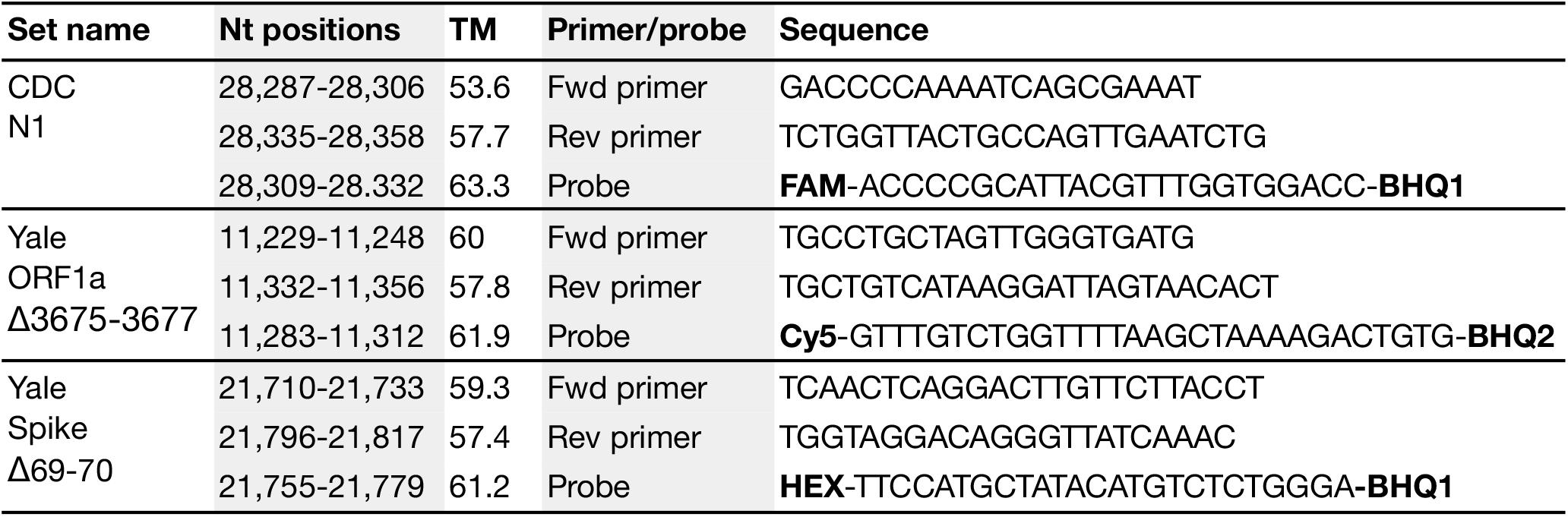
Primers and probes used in the multiplexed RT-qPCR variant screening assay.

**Supplementary Table 3:** Interpretation of results from the multiplexed RT-qPCR variant screening assay.

| Result | N1-FAM | Orf1a-Cy5 | Spike-HEX |
| --- | --- | --- | --- |
| Potentially B.1.1.7 (VOC) | CT ≤ 35 | Undetected | Undetected |
| Potentially B.1.351 or P.1 (VOC) | CT ≤ 35 | Undetected | CT ≤ 35 |
| Potentially B.1.375 (not a VOC) | CT ≤ 35 | CT ≤ 35 | Undetected |
| Other lineages | CT ≤ 35 | CT ≤ 35 | CT ≤ 35 |
| Inconclusive | CT > 35 or undetected | Any value | Any value |

VOC, variant of concern. Thresholds need to be determined when using different RT-qPCR reagents or instruments.

